# Dissecting the Vascular-Cognitive Nexus: Energetic vs. Conventional Hemodynamic Parameters

**DOI:** 10.1101/2023.11.06.23298188

**Authors:** Hao-Min Cheng, Jiun-Jr Wang, Shao-Yuan Chuang, Chen-Hua Lin, Gary F. Mitchell, Chi-Jung Huang, Pei-Ning Wang, Chih-Ping Chung, Liang-Kung Chen, Wen-Harn Pan, Li-Ning Peng, Chen-Huan Chen

**Author notes:** Correspondence to: Dr. Chen-Huan Chen, Address: No. 155 Li-Long St., Sec. 2, Beitou District, Taipei, Taiwan. contribute equally to the present work (Jiun-Jr Wang, Li-Ning Peng). **Disclosures:** G.F.M. is owner of Cardiovascular Engineering, Inc., a company that designs and manufactures devices that measure vascular stiffness. The company uses these devices in clinical trials that evaluate the effects of diseases and interventions on vascular stiffness. G.F.M. also serves as a consultant to and receives grants and honoraria from Novartis, Merck, Bayer, Servier, Philips, and deCODE genetics and is an inventor on a pending patent application that discloses a method for estimating carotid-femoral pulse wave velocity and vascular age by using a convolutional neural network. The remaining authors declare that they have no competing interests.

## Abstract

**Background:** Blood flow measurements are being studied in relation to vascular health and cognitive function, but their role is unclear.

**Objective:** We investigated whether energetic hemodynamic parameters, such as aortic and carotid mean and pulsatile energy, and energy pulsatility index (PI), provide a more nuanced understanding of the vascular-cognitive link, as assessed by the Montreal Cognitive Assessment (MoCA), than conventional flow and flow PI.

**Methods:** Cognitive evaluation and hemodynamic measurements, including aortic and carotid pressure and flow waves, were performed on 1858 MoCA participants. Energy was calculated by integrating pressure time flow. An asymmetric bifurcation model was used to calculate aortic and carotid mean, pulsatile energy, and hemodynamic parameters across the interface.

**Results:** After adjusting for age, sex, education, depression score, heart rate, BMI, HDL-cholesterol, and glucose levels, energetic hemodynamic parameters were more associated with MoCA score than aortic and carotid flow and flow PI. In particular, carotid mean energy was most significantly positively associated with MoCA (standardized beta = 0.053, P = 0.0253) and energy PI was most significantly negatively associated (standardized beta = -0.093, P = 0.0002), surpassing conventional metrics like carotid PI. Aortic pressure reflection coefficient at the aorta-carotid bifurcation was positively correlated with mean carotid energy and weakly negatively correlated with PI. Aortic characteristic impedance positively correlated with carotid energy PI but not mean energy.

**Conclusion:** Our study shows that energetic hemodynamic parameters, particularly carotid mean energy and energy PI, better explain the vascular-cognitive nexus than conventional measures.

## Background

Cognitive decline and dementia are escalating global health concerns, especially within aging populations ^1, 2^ A robust body of evidence links vascular dementia and Alzheimer’s disease with hemodynamic abnormalities stemming from various cardiovascular conditions.^3,4^. High blood pressure, for example, compromises the cerebral vasculature and predisposes individuals to cognitive impairment, especially when it develops earlier in life. ^5–8^

Traditionally, carotid hemodynamics have been a focus of study given their role in cerebral blood supply. Altered carotid pressure and flow waveforms have been implicated in cognitive dysfunction, yet the mechanisms remain inadequately understood.^9–12^ We previously demonstrated that lower carotid flow velocity was frequently associated with smaller cerebral white matter and gray matter volume,^10^ and has been regarded as a strong indicator of brain atrophy and an effective predictor of increased risk of stroke.^9, 13^ Previous research has emphasized parameters like pressure and flow pulsatility indexes (PIs) for their associations with cerebrovascular diseases and cognitive decline.^13, 14^

Traditional metrics, which primarily concentrate on the dynamics of pressure and flow, might not comprehensively capture the complex interaction between vascular and cognitive functions. Our research argues that energetic hemodynamic parameters provide a comprehensive and nuanced framework for analyzing this complex relationship, surpassing the explanatory capability of conventional pressure and flow indices when placed side by side.^15^

The “impedance mismatch” hypothesis has been proposed to illustrate vascular aging-related target organ damage. The hypothesis states that pulsatile energy is not fully transmitted into the distal vasculature because impedance mismatch at the junction of the highly compliant aorta and relatively stiff first generation branch vessels limits transmission of pulsatile power/energy into the carotid artery.^15^ Vascular aging of proximal aorta erodes impedance mismatch and permits transmission of excess pulsatile energy into the carotid arteries. Although the impedance mismatch hypothesis is based on sound physical principles, few studies have examined the hypothesis.

In order to bridge these existing knowledge gaps, our research study utilizes a sample of 1858 participants from two distinct cohorts. We employ an asymmetric arterial bifurcation model to obtain a comprehensive collection of hemodynamic measures.^16^ These measures encompass not only conventional metrics based on pressure and flow, but also energetic parameters as depicted in Figure 1. Our objective is to control for potential confounding variables in order to assess the relative effectiveness of these parameter in elucidating relations of hemodynamic measures with cognitive function, as well as to evaluate the validity of the impedance mismatch hypothesis.

**Figure 1.**
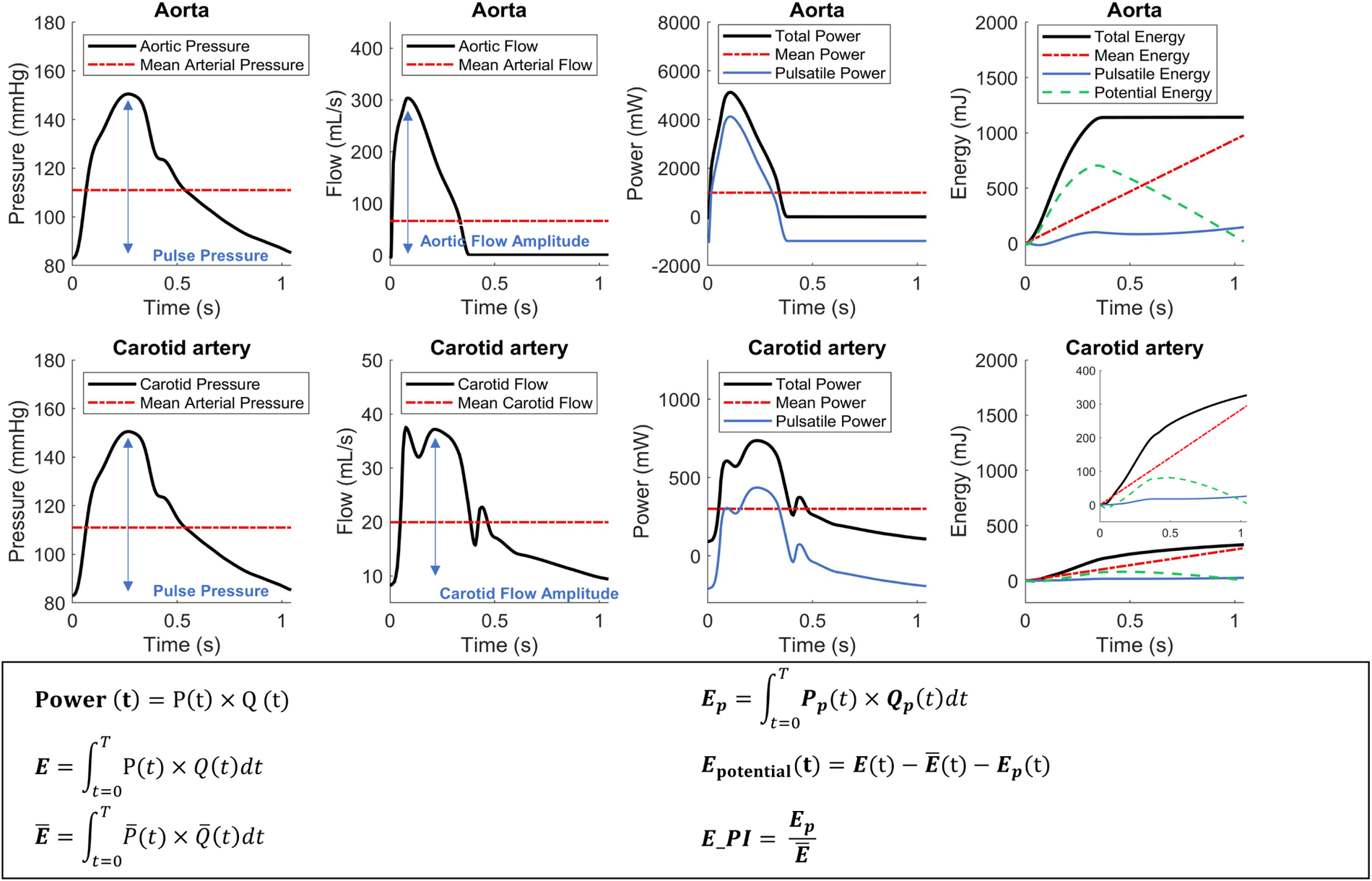
The presentation of the hydraulic total, mean, pulsatile and potential energy (the 1^st^ right column), calculated at the central aorta (top panel) and carotid artery (bottom panel), as a function of time. Three steps were required to conclude the energy calculation. The first step was to calculate the total, mean and pulsatile power as a function of time (the 2nd right column). The total power (black curve) was calculated as the product of concurrent aortic pressure (black curve, the 1st left column) and aortic flow (black curve, the 2nd left column). Similarly, the mean power (red dash-dot curve) and pulsatile power (blue solid curve) were calculated as the product of concurrent mean pressure and mean flow (red dash-dot curve) and concurrent pulsatile pressure and pulsatile flow, respectively. The second step was to calculate the total, mean and pulsatile energy, by integrating the total, mean and pulsatile power with respect to time (the 1st right column). The third step, the potential energy as a function of time (green dashed curve) was calculated as the instantaneous difference between the total energy (black curve) and the sum of the mean (red dash-dot curve) and pulsatile energy (blue solid curve; the 1st right column). Note that the total energy was not equal to mean energy plus pulsatile energy until the end of a cycle.

## Materials and Methods

### Study cohorts

The present study cohort comprised two study populations, the Cardiovascular and Disease Risk Factors Two-Township Study (CVDFACTS) and the Longitudinal Aging Study of Taipei (LAST). CVDFACTS is an ongoing longitudinal study of the risk factors and pathogenesis of cardiovascular disease in two Taiwanese cities, Chu-Dung (a Hakka community) and Pu-Tzu (a Fukienese community).^17^ Residents who were aged 30 and over and previously participated in one or more of CVDFACTS surveys were recruited from 2017 through 2020. The Longitudinal Aging Study of Taipei (LAST) is an ongoing community-based study that was initiated by Aging and Health Research Center of the National Yang Ming Chiao Tung University from May 2016 to December 2019, a total of 1532 community volunteers were invited to participate the first wave of the study. For the cardiovascular hemodynamic assessments, these two study cohorts have adopted the same study protocol, which has been approved by the Institutional Review Board of National Yang Ming Chiao Tung University. Each participant was well-informed, and a written consent was obtained before the study.

All subjects were scheduled for two visits within three months for the study. Information of personal characteristics, prior medical history, anthropometric measurements, cognitive function, and fasting blood tests were collected during the first visit. Medical history, particularly stroke and heart diseases, was acquired by structured questionnaires. An example was as follows: “Did you have heart disease diagnosed by a physician at a clinic or hospital?” Cardiovascular hemodynamic measurements were conducted during the second visit. We used structured questionnaires, the Center for Epidemiologic Studies Depression Scale (CESD) and Taiwanese Depression Scale (TDS) in LAST cohort and CVDFACTS cohort, respectively, to measure depression. The depression scales of these two cohorts were then normalized for further analysis.

#### Cognitive function

The global cognitive function was evaluated using the Montreal Cognitive Assessment (MoCA) protocol with the Chinese version specifically used in Taiwan,^18^ through face-to-face interview by dedicated and qualified nurses adherent to the standardized study guide. The MoCA was constituted by 20 items clustered into 7 subgroups, each dedicated to one aspect of cognitive function, namely executive function/visuospatial ability (5 points), attention (6 points), animal naming (3 points), language (3 points), abstraction (2 points), short-term memory (5 points), and orientation (6 points) with a total score of 30 points.^19^

#### Echocardiography

Participants all received transthoracic echocardiography performed by an experienced sonographer. All images were acquired using a commercially available machine (HD11 XE Ultrasound system, Koninklijke Philips N.V.) and digitized using the TomTec Image-Arena™ Software 4.0 (TomTec Imaging Systems GmbH, Munich, Germany) by the same sonographer. Left ventricular (LV) volume was acquired by tracing the endocardial border of the left ventricle at both the end-diastole and end-systole, then summing up a stack of elliptical disks in apical 4-chamber view. The determination of left ventricular ejection fraction involved calculating the discrepancy between the volume of the left ventricle at the end of diastole and the volume at the end of systole. Doppler-derived stroke volume was the product of the cross-sectional area and the velocity, calculated via Doppler signal acquired at the LV outflow tract during systole.^20^ Doppler-derived cardiac output was calculated as the product of stroke volume and heart rate. Cardiac index (CI) was calculated as cardiac output divided by the body surface area.^20^

#### Arterial stiffness

Arterial waveforms at the right common carotid artery and the right femoral artery were recorded in sequence, by means of applanation tonometry using a pencil-type tonometer, a high-fidelity strain-gauge transducer at the flat tip of 7-mm-diameter (SPC-350, Millar Instruments Inc, Texas).^21^ Body surface measurements from carotid to femoral pulse recording sites were obtained by tape measure. Carotid-femoral pulse wave velocity (cf-PWV) was calculated as the distance between the two measurement sites, divided by the foot-to-foot wave transit time. Transit time was calibrated by the simultaneously recorded ECG, and aligned via a custom-designed software on a commercial software package (Matlab, version 4.2, The MathWorks, Inc.).^21^

### Data Acquisition and Analysis

Each recording lasted for 25 seconds (5–6 respiratory cycles) to ensure high-quality pulse waveforms. Each waveform admitted for subsequent hemodynamic analysis was an average of 10 consecutive steady waveforms. The aortic pressure was determined by the tonometry waveform measured at the right common carotid artery (CCA), calibrated by mean and diastolic pressure of the brachial cuff pressure. Central aortic blood flow was determined as the Doppler flow velocity measured using a pulsed-wave Doppler echocardiography at the LV outflow tract in an apical five-chamber view, multiplied by the cross-sectional area at the LV outflow tract on a parasternal long-axis views. The left and right common carotid flows were determined by the Doppler velocity waveform, recorded using a linear array probe with 3.1–10.0 MHz imaging frequency in the longitudinal view. The sample volume was placed at the center of the CCA around 1 cm proximal to the carotid bulb. Flow velocities were multiplied by the respective carotid lumen cross-sectional area to get volumetric flow rate. The carotid arterial diameter was measured from the intima-lumen interface of the near wall to the lumen-intima interface of the far wall. All Doppler measurements were obtained with an insonation angle maintained ≤60°. The carotid flow waveform images were digitized and transformed into a signal-averaged flow spectrum in the MATLAB program. Each velocity waveform admitted for subsequent analysis was an average of 10 consecutive waveforms. Since the pressure at ascending aorta is largely comparable to the pressure at carotid artery, the measured carotid arterial pressure waveform was adopted as the aortic pressure waveform to be paired with the corresponding aortic flow for hemodynamic analysis. Given that the passage of a wave causes simultaneous changes in both pressure and flow, arterial pressure was shifted in time so that the onset of systolic pressure matches that of the blood flow. For carotid hemodynamic analysis, the carotid blood flow admitted for hemodynamic analysis was the sum of both the left and right carotid blood flows.

### Hemodynamic analysis

#### The mean and pulsatile hydraulic energy

In this study, we calculated the hydraulic mean, pulsatile energy and total energy for one cardiac cycle, consistent with those power-based parameters adopted by Haidar et al.^16^ To calculate the hydraulic mean and pulsatile energy for one cardiac cycle, we first separated the measured pressure *P*(*t*) and blood flow *Q*(*t*) waveforms into their respective mean and pulsatile components:

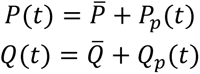

Total and pulsatile hydraulic energy, *E,* of one cardiac cycle, *T,* was calculated as the product of pressure *P*(*t*) and flow *Q*(*t*) or pulsatile pressure *P*_*p*_(*t*) and flow *Q*_*p*_(*t*), respectively, integrated with respect to time *t* over *T*. Calculations were repeated for both aorta and carotid artery. In addition, we computed an effective accumulating “mean” energy, as *P̄* × *Q̄* × *t* (**Figure 1** **and Supplemental Table 1**).

### Aortic and carotid wave pressure, flow and energy reflection and transmission coefficients

A simplified bifurcation model was adopted to interpret hemodynamics at the interface of the aortic-carotid bifurcation,^16^ which was presumed as the central aorta branching into two asymmetric daughter vessels, a larger downstream thoracic aorta and a smaller carotid artery. In this model, the carotid blood flow was the sum of both the left and right carotid flow.^16^

The aortic forward-going wave (defined as the direction of the mean blood flow) pressure reflection coefficient at the aortic-carotid bifurcation, Γ_*Ao*_, was calculated as

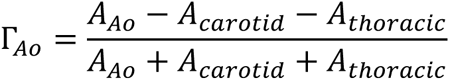

where *A_*Ao*_*, *A*_*carotid*_, *A*_*thoracic*_ were the admittance of the proximal aorta, the carotid artery and the downstream thoracic aorta, respectively. The admittance is the reciprocal of impedance. At aorta and carotid artery, admittance was calculated as the averaged ratio of magnitudes of flow to magnitudes of pressure from the 2^nd^ harmonic through the 10^th^ harmonic in frequency domain. At thoracic aorta, admittance was calculated as *A*_*Ao*_ multiplied by the ratio of thoracic mean flow, which was the difference between the aortic mean flow and the carotid mean flow, divided by aortic mean flow, following the assumption that the local pulse wave velocity and mean flow velocity were presumed uniform across aorta just proximal of the bifurcation junction and distal thoracic artery^15^. By definition, the aggregate forward-going flow wave reflection coefficient at the aortic-carotid bifurcation was −Γ_*Ao*_.

### Statistical methods

Continuous and categorical characteristics of the sample were described by mean (± standard deviation) and percentage, respectively. For the baseline model, the partial correlation coefficients between MoCA score and aorta-carotid arterial hemodynamics were computed by adjusting for age, sex, education, depression score and heart rate. For the fully adjusted model, we further adjusted for other potential confounders including body mass index, and LDL-cholesterol, and fasting glucose, for the associations between MoCA score and aorta-carotid arterial hemodynamics in addition to the baseline model.

The Pearson correlation matrix was conducted for the interrelations of carotid energy PI, carotid flow PI, aortic energy PI, aortic pressure PI and aortic flow PI. Correlations between carotid total, mean and pulsatile energy with aortic total, mean and pulsatile energy, aortic forward wave pulsatile energy transmitted into carotid artery, and Γ_*Ao*_ were examined by using the Pearson correlation. The correlates of carotid mean and pulsatile energy, and carotid energy PI were identified by general linear models.

We employed causal mediation models to examine the relations between Zao and cognitive function. Additionally, we investigated whether these associations were mediated by the carotid energy pulsatility index and carotid mean energy, or by the carotid flow pulsatility index and carotid mean flow. Furthermore, we explored the associations between aortic pressure wave reflection coefficient (Γ_Ao) and cognitive function. We also examined whether these associations were mediated by the carotid energy pulsatility index and carotid mean energy, or by the carotid flow pulsatility index and carotid mean flow. The statistic significant p-value was dependent on the multiple comparison testing.

The Path analysis, involving causal mediation models, was conducted using the CALIS procedure in SAS 9.4. The performance of all models was assessed using the goodness of fit index (GFI). The significance level was established at a value of 0.05.

## Results

Clinical characteristics and hemodynamic parameters of all 1858 participants are summarized in Table 1. Females have a higher average age, BMI, and HDL-cholesterol than males. Males report hypertension and diabetes more than females. Males have more university degrees than females. Men and women have a median Montreal cognitive score of 27.

**Table 1.** Characteristics of study population.

| Variables (mean ± std. / %) | Females | | Males | |
| --- | --- | --- | --- | --- |
| *N* | **1183** |  | **675** |  |
| Age, years | 61.2 | 9.9 | 59.4 | 11.4 |
| Body mass index, kg/m<sup>2</sup> | 23.5 | 3.4 | 24.8 | 3.1 |
| Waist circumference, cm | 79.1 | 9.1 | 86.9 | 8.4 |
| Triglycerides, mg/dl | 114.6 | 78.7 | 129.1 | 93.6 |
| HDL-cholesterol, mg/dl | 63.5 | 16.1 | 51.1 | 12.3 |
| LDL-cholesterol, mg/dl | 120.3 | 31.8 | 116.0 | 35.0 |
| Total cholesterol, mg/dl | 208.2 | 35.6 | 193.1 | 38.2 |
| Fasting glucose, mg/dl | 95.5 | 17.5 | 100.1 | 19.1 |
| Medical history, n (%) |  |  |  |  |
| hypertension, % | **26.54%** |  | **36.15%** |  |
| hypertensive medicine, % | **15.64%** |  | **15.85%** |  |
| diabetes, % | **8.71%** |  | **13.78%** |  |
| anti-diabetic medicine, % | **7.44%** |  | **11.56%** |  |
| Education level, n (%) |  |  |  |  |
| Elementary or below | **8.96%** |  | **2.22%** |  |
| Junior school | **9.97%** |  | **7.70%** |  |
| High school | **34.23%** |  | **25.48%** |  |
| University of higher | **46.83%** |  | **64.59%** |  |
| Brachial systolic blood pressure, mmHg | 123.1 | 17.7 | 128.8 | 16.7 |
| Brachial diastolic blood pressure, mmHg | 72.5 | 9.3 | 78.9 | 9.8 |
| Carotid-femoral pulse wave velocity, m/sec | 12.0 | 3.3 | 12.6 | 4.0 |
| $Z_{ao}$, dyne*s/cm<sup>5</sup> | 126.0 | 61.7 | 122.4 | 63.1 |
| Aortic pressure wave reflection coefficient, $\Gamma_{Ao}$ | 0.11 | 0.06 | 0.10 | 0.06 |
| Montreal cognitive assessment score, median (25%, 75%) | **27 [26, 29]** |  | **27 [26, 29]** |  |

HDL = high density lipoprotein; LDL = low density lipoprotein; $Z_{ao}$ = aortic characteristic impedance.

The carotid mean flow was found to be approximately 30.3% of the aortic mean flow. Additionally, the carotid pulsatile flow was observed to be around 10.2% of the aortic pulsatile flow. Consequently, the carotid flow pulsatility index (PI) was calculated to be approximately 34.5% of the aortic flow PI. The energy pulsatility index (PI) of the carotid artery was found to be 37.9% when compared to the aortic counterparts, as indicated in Table 2. A comparative analysis of flow and energy metrics between women and men at ascending aorta and common carotid artery is summarized in Table 2. As expected, men exhibited higher mean and peak flow rate and total energy at both ascending aorta and carotid artery. Still, there were no significant differences between men and women in terms of energy pulsatility at both sites.

**Table 2.**
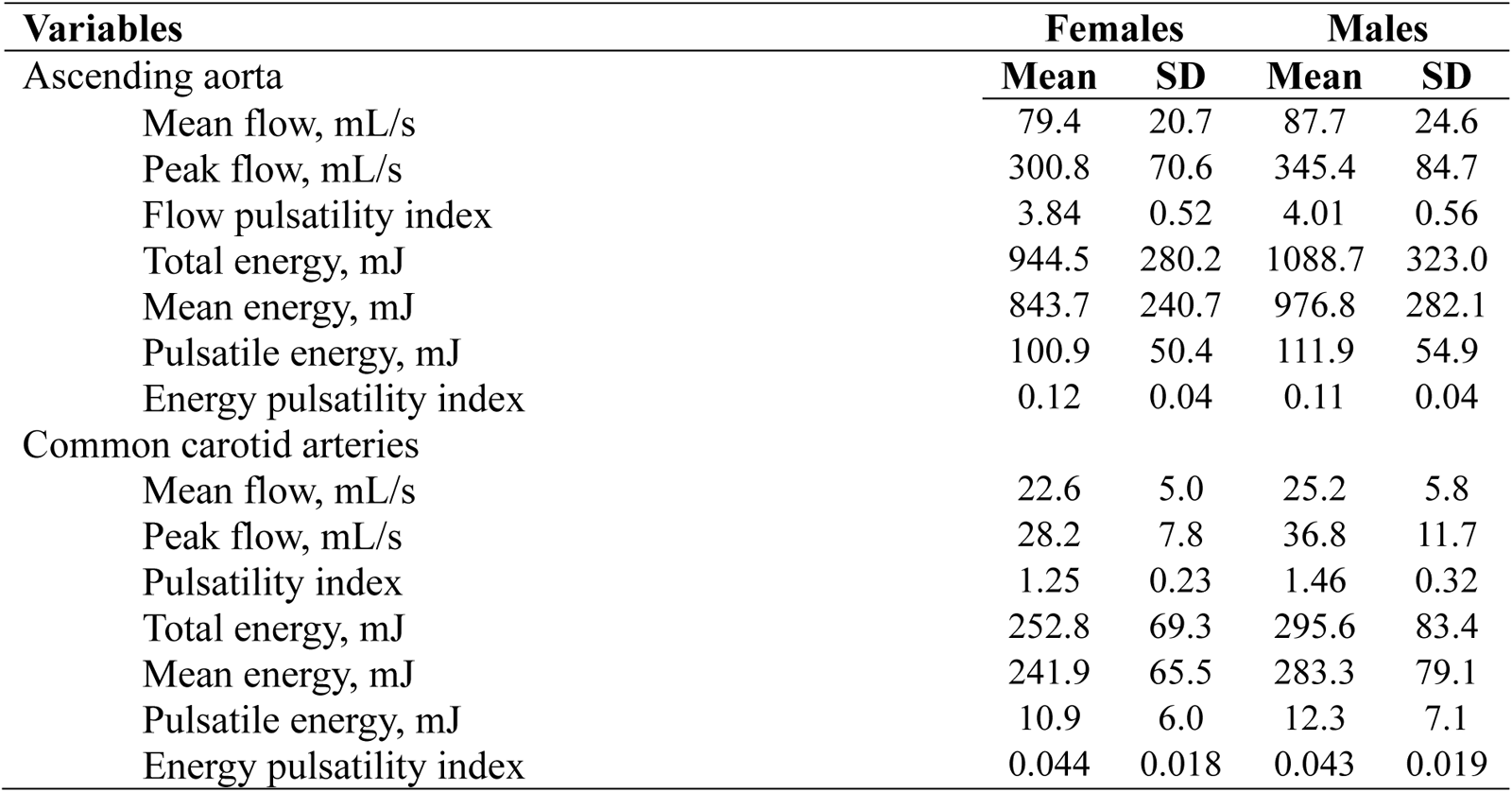
Flow and energy related variables at ascending aorta and common carotid arteries.

The MoCA score demonstrated significant associations with variables such as age, education level, blood pressure, fasting glucose levels, and lipid profiles, as indicated in Supplemental Table 2. After adjusting for age, sex, educational attainment, and depression levels, only certain variables, namely peripheral mean pressure, waist circumference, body mass index, high density lipoprotein cholesterol, total cholesterol, and fasting blood sugar, continued to exhibit a significant association with MoCA. The associations between MoCA score and different hemodynamic parameters are displayed in Table 3. The findings indicate that there are stronger associations between carotid hemodynamic parameters and MoCA scores compared to aortic hemodynamic parameters. Specifically, carotid mean energy, pulsatile energy, and energy pulsatility index exhibit significantly stronger associations than metrics based on pressure or flow, as shown in Table 4. The MoCA scores demonstrated inverse relationships with a number of parameters, such as the aortic pressure, carotid, and aortic energy PIs, and carotid flow PI. However, they exhibited a positive correlation with the carotid mean energy. Despite controlling for variables such as age, gender, level of education, and health indicators, the majority of these associations continued to hold. It is worth noting that the carotid energy PI exhibited the strongest correlation with MoCA scores.

**Table 3.** Associations of flow and energy-related variables and carotid-femoral pulse wave velocity with MoCA score.

| Variables | Crude |  | Model A |  | Model B |  |
| --- | --- | --- | --- | --- | --- | --- |
| | $\beta$ | P value | $\beta$ | P value | $\beta$ | P value |
| Ascending aorta |  |  |  |  |  |  |
| Mean flow, mL/s | -0.040 | 0.0833 | -0.030 | 0.1966 | -0.017 | 0.4788 |
| Peak flow, mL/s | -0.026 | 0.2540 | -0.012 | 0.5911 | -0.002 | 0.9364 |
| Flow pulsatility index | 0.013 | 0.5713 | 0.032 | 0.2353 | 0.0232 | 0.3916 |
| Total energy, mJ | **-0.072** | **0.0020\*** | -0.021 | 0.3362 | -0.002 | 0.9367 |
| Mean energy, mJ | **-0.056** | **0.0153** | -0.010 | 0.6366 | 0.009 | 0.7000 |
| Pulsatile energy, mJ | **-0.132** | **<0.0001\*** | **-0.071** | **0.0014\*** | **-0.054** | **0.0185** |
| Energy pulsatility index | **-0.141** | **<0.0001\*** | **-0.088** | **<0.0001\*** | **-0.076** | **0.0006\*** |
| Common carotid arteries |  |  |  |  |  |  |
| Mean flow, mL/s | **0.083** | **0.0003\*** | 0.043 | 0.0529 | 0.038 | 0.0856 |
| Peak flow, mL/s | 0.039 | 0.0901 | -0.021 | 0.3677 | -0.013 | 0.5880 |
| Pulsatility index | **-0.047** | **0.0435** | **-0.084** | **0.0002\*** | **-0.069** | **0.0022\*** |
| Total energy, mJ | 0.022 | 0.3412 | **0.047** | **0.0484** | **0.053** | **0.0253** |
| Mean energy, mJ | 0.0349 | 0.1323 | **0.0529** | **0.0248** | **0.058** | **0.0136** |
| Pulsatile energy, mJ | **-0.133** | **<0.0001\*** | -0.047 | 0.0524 | -0.033 | 0.1805 |
| Energy pulsatility index | **-0.198** | **<0.0001\*** | **-0.1074** | **<0.0001\*** | **-0.093** | **0.0002\*** |
| Carotid-femoral pulse wave velocity, m/sec | **-0.175** | **<0.0001\*** | -0.0084 | 0.7545 | 0.00622 | 0.8182 |

\*:p-value<0.0033 (0.05/15) for multiple comparison.
$\beta$ : standardized $\beta$ regression coefficient
Model A: adjusted for age, sex, education, depression, and heart rate
Model B: adjusted for age, sex, education, depression, heart rate, body mass index, high density lipoprotein-cholesterol and glucose.

**Table 4.**
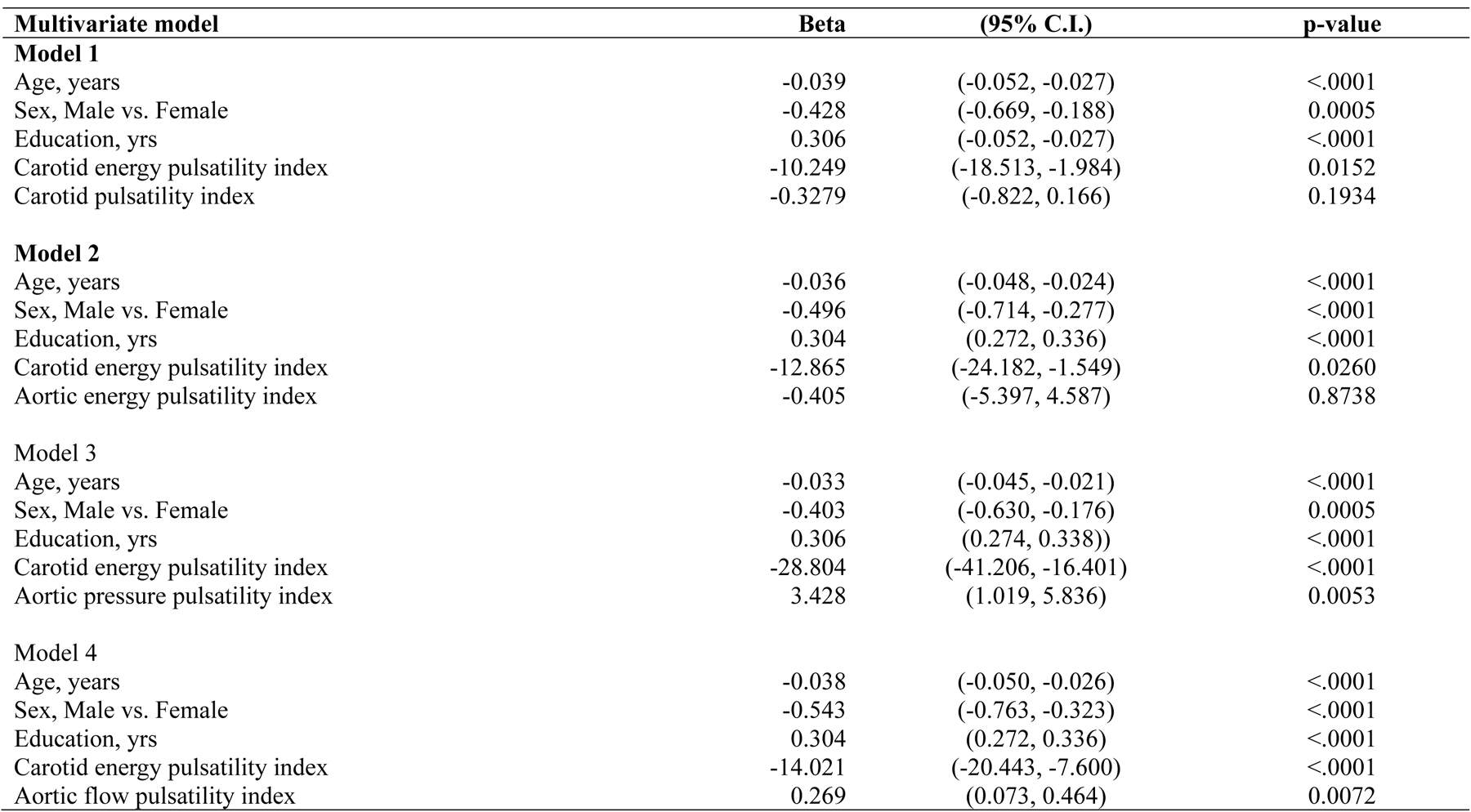
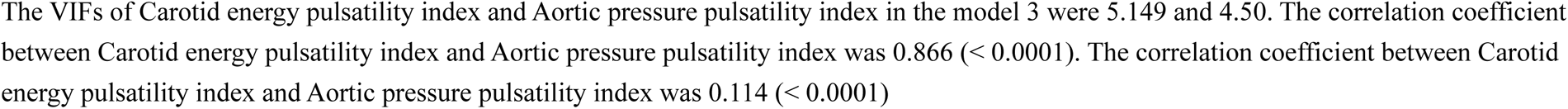
The comparative associations of carotid energy pulsatility index, carotid flow pulsatility index, aortic energy pulsatility index with M_O_CA scores.

Subsequent examination indicated that the cognitive function exhibited a stronger correlation with the carotid hemodynamic pulsatility indices compared to the central aortic hemodynamic pulsatility indices. Among the various hemodynamic indices related to the aortic and carotid arteries, it was observed that the carotid energy PI exhibited the most significant correlation with cognitive function (r= -0.097). This was followed by the carotid flow PI (r=-0.085), and the aortic energy PI (r = -0.080), as depicted in Figure 3A.

In subsequent multivariable analyses, after adjusting for age, sex, and education, it was found that carotid energy PI remained significantly associated with cognitive function. However, carotid flow PI (Model 1 in Table 4) and aortic energy PI (Model 2 in Table 4) were no longer significantly associated with cognitive function. These findings support the notion that carotid energy PI, an energy-based hemodynamic parameter, holds superior prognostic significance and is the most effective contributor to cognitive function.

Table 5 provides a comprehensive analysis of relations between aortic flow metrics and carotid energy indicators and flow PI. There is a positive correlation between the carotid mean energy and both the aortic mean flow (r = 0.126, p-value < 0.0001) and peak flow (r = 0.11, p-value < 0.0001). Similarly, it can be observed that the carotid energy PI exhibits a positive correlation with aortic mean flow (r = 0.104), as well as a weaker correlation with peak flow (r = 0.082). The carotid pulsatility index is noteworthy for its robust positive associations with various aortic measures, particularly with peak flow (r = 0.186).

**Table 5.**
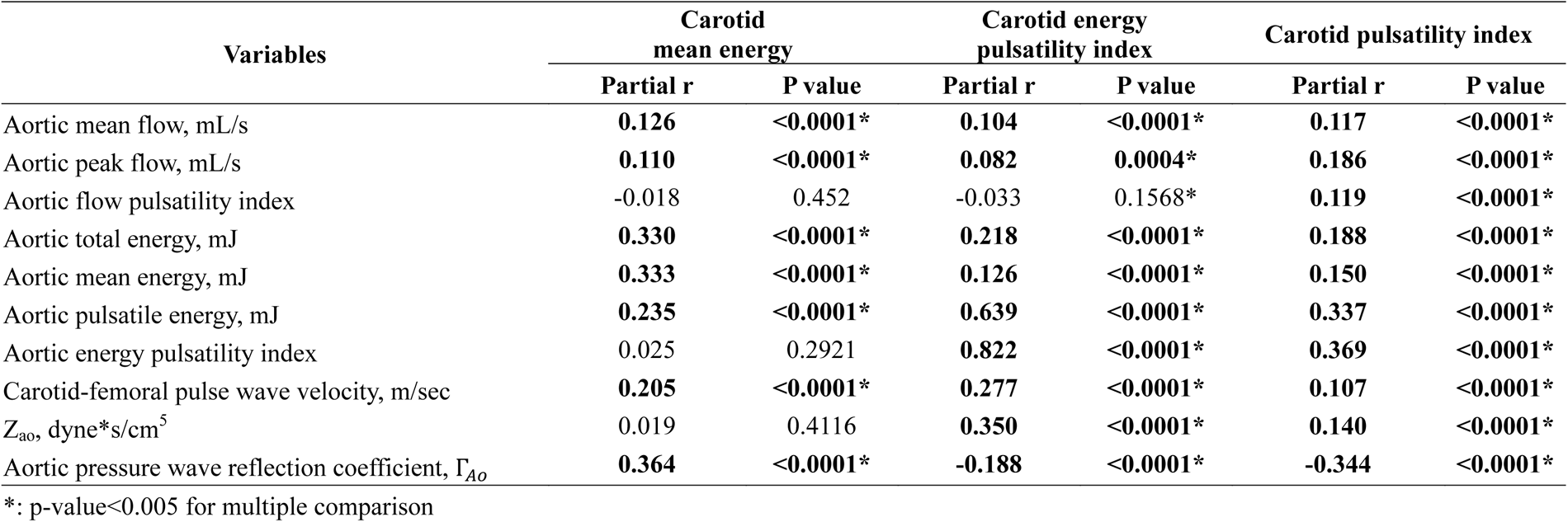
Partial correlation coefficients, adjusted for age, sex and heart rate.

Based on the findings presented in Table 5, a significant positive correlation was identified between aortic characteristic impedance and both carotid energy PI and carotid PI. It is noteworthy that all p-values associated with these correlations were found to be less than 0.0001. It is important to emphasize the presence of a correlation between the aorta-carotid reflection coefficient, Γ_Ao, and specific hemodynamic parameters. More specifically, as the value of Γ_Ao increases, there is a corresponding decrease in carotid energy PI and carotid flow PI. However, carotid mean energy shows a positive correlation with Γ_Ao.

The analysis presented in Figure 2 clearly demonstrates the impact of carotid energy pulsatility index and carotid mean energy on MoCA scores. The impact of Zao and the wave reflection coefficient on MoCA is solely mediated through energy metrics, without any direct influence. On the other hand, the flow-based analysis places emphasis on the contribution of flow pulsatility to the MoCA, ascribing significant direct effects to both Zao (with borderline significance) and the wave reflection coefficient.

**Figure 2.**
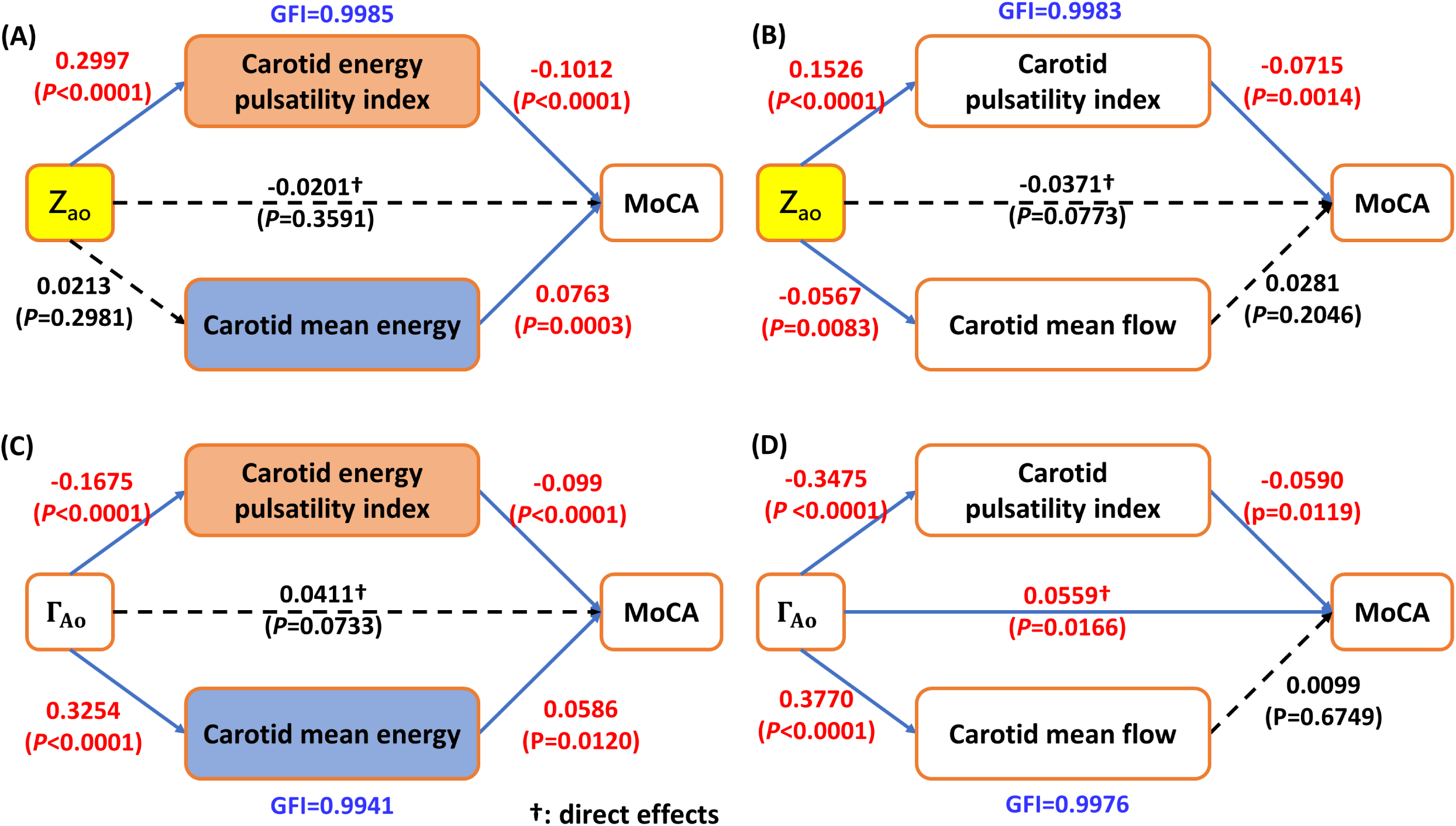
Analysis of the causal mediation of the associations between Zao and Aortic pressure wave reflection coefficient and cognitive function. (A) Zao was indirectly associated with cognitive function through carotid energy pulsatility index, but not by carotid mean energy. Zao did not have direct effect on cognitive function (beta=-0.0201, p=0.3591). (B) Zao was indirectly associated with cognitive function through carotid energy pulsatility index, but not by carotid mean energy. Carotid mean flow was not associated with cognitive function. (C) Aortic pressure wave reflection coefficient (Γ_Ao) was associated with cognitive function through carotid energy pulsatility index and aortic pressure wave reflection coefficient (Γ_Ao) had not direct effect on cognitive function. (D) Aortic pressure wave reflection coefficient (Γ_Ao) was associated with cognitive function through carotid energy pulsatility index and aortic pressure wave reflection coefficient (Γ_Ao) also had a direct effect on cognitive function (beta = 0.0559, p=0.0166).

**Figure 3.**
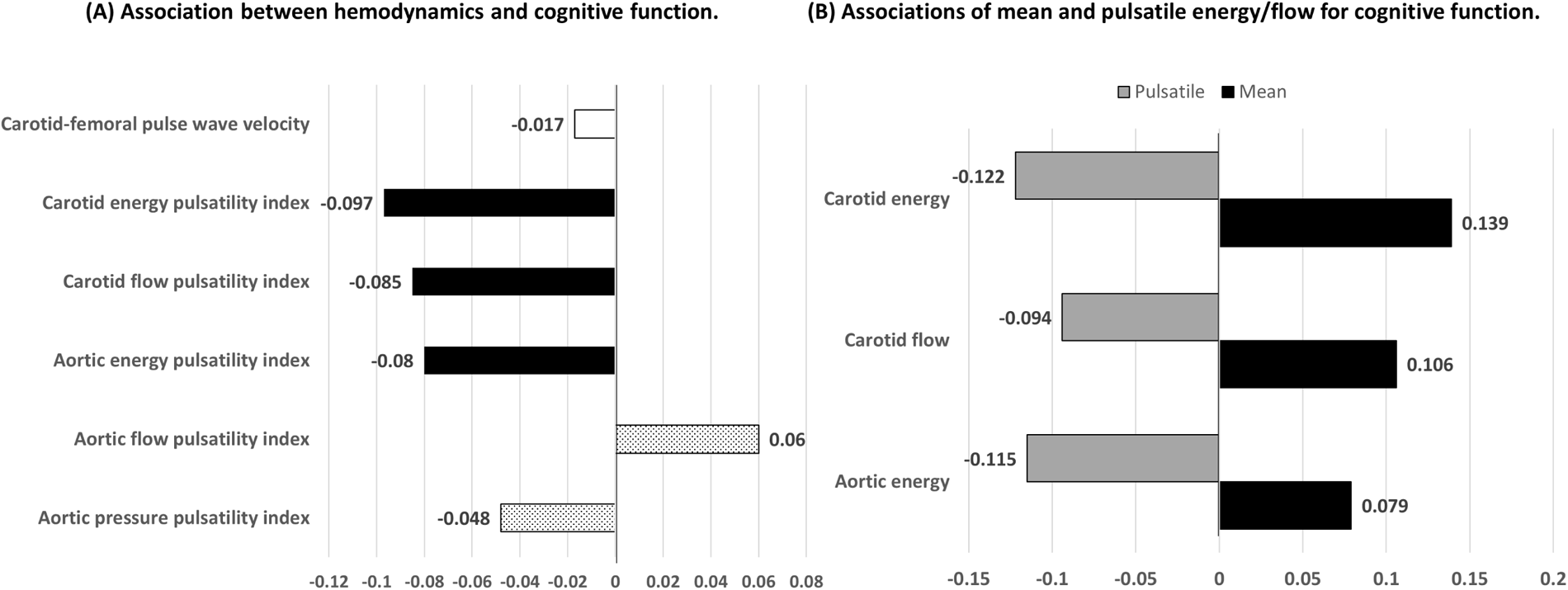
Differential aortic or carotid hemodynamics in relation to cognitive function. (A) The correlation between hemodynamics and cognitive function; (B) The correlations between cognitive function and mean energy and pulsatile energy. Coefficients of correlation were all adjusted for sex, age, and level of education. The histogram in black in Figure (A) represents a p-value less than 0.01, the dot histogram a p-value between 0.01 and 0.05, and the white histogram a p-value greater than 0.05. Standardized beta of hemodynamics and MOCA in multivariable models, with age, gender, and education adjustments, are displayed in Figure (B). Every standardized beta value attained statistical significance (p < 0.05).

## Discussion

This study emphasizes the unique importance of energetic parameters in comprehending the connection between vascular health and cognitive function. The initial analysis revealed a clear positive relationship between carotid mean energy and cognitive function, while a negative relationship was observed with pulsatile energy. Our research represents the initial effort in identifying the carotid energy PI as a more effective hemodynamic indicator for cognitive function. Furthermore, after conducting a thorough evaluation of numerous hemodynamic parameters related to the aorta and carotid artery, it has been established that the carotid energy PI emerges as the primary predictor of cognitive performance, surpassing the predictive ability of previously proposed pressure and flow-based metrics. We subsequently conducted an investigation into the carotid energy pulsatility index (PI) and carotid mean energy, in relation to aortic flow and energy indices, aortic stiffness, aortic-carotid impedance mismatch, and organ perfusion.

The aortic-carotid impedance mismatch hypothesis has long been regarded playing a critical role in cognitive function. We showed that an increase in proximal aortic stiffness is a contributing factor to the elevated pulsatile energy observed in the carotid arteries, whereas the influence of aortic stiffness, Zao, and the wave reflection coefficient, Γ_Ao, on MoCA predominantly manifests itself through energy metrics, rather than through direct effects. In contrast, a flow-based assessment underscored the fundamental importance of flow pulsatility in the Montreal Cognitive Assessment (MoCA). Significant direct effects were ascribed to both Zao and the wave reflection coefficient, Γ_Ao. However, it is conceivable that this assessment may have placed excessive emphasis on the importance of wave reflection in the pathophysiological connection between vascular health and cognitive impairment. Hence, our research approach, which is based on the concept of energy, focuses on the complex impact of energy pulsations on cognitive function. This approach contributes to a comprehensive understanding of the interconnected relationship between vascular health and cognitive function.

Our research comprehensively elucidates the fundamental mechanisms that govern the transfer of increased circulatory pulsatility from a stiffened aorta to cerebral circulation, confirming the significant impact of the “impedance mismatch” phenomenon. As individuals grow older, there is a noticeable increase in impedance “matching,” which can be attributed to a greater increase in aortic impedance as compared to impedance of first-generation branch vessels. This impedance matching leads to heightened transmission of pressure and flow pulsatility into the cerebral circulation, ultimately strengthening the connection between the cardiac and cerebral structures and their respective functions.^15^ These observations align with the gradual stiffening of the proximal aorta, as depicted in Figure S1. Furthermore, our analysis comprehensively investigates the substantial impacts of the carotid energy pulsatility index and carotid mean energy on MoCA scores.

This observation implies that the flow-based approach may potentially exaggerate the extent to which wave reflection contributes to the preservation of pulsatility, while simultaneously overlooking its influence in amplifying mean energy. By employing the energy-based methodology, it is possible to distinguish the heightened influence of energy pulsatility compared to mean energy on the Montreal Cognitive Assessment (MoCA), with Zao playing a noteworthy role. Nevertheless, the contributions of protective effects resulting from reflections at the interface between the aorta and carotid artery are somewhat limited.

Our finding was consistent with the AGES-Reykjavik Study, where the pulsatile power (pulsatile energy normalized by the period of a cardiac cycle) was negatively associated with cognitive function.^16^ In the present study, we further demonstrated the counteractive effect of the positive association between carotid mean energy and cognitive function was comparable to the negative association between carotid pulsatile energy and MOCA score (standardized Beta: 0.117 vs. -0.109).

### Study strength and limitations

There are several strengths in this study. First, we showed that the carotid mean energy and the pulsatile energy were positively and negatively associated with cognitive function, respectively. Considering that this is the first study to identify the carotid energy PI as the best hemodynamic indicator for cognitive function, our study findings should be reproduced by other studies. Second, through extensive evaluation of exhaustive aorta-carotid hemodynamic parameters, we concluded that the carotid energy PI is the single most effective hemodynamic parameter for predicting cognitive function, much more effective than any other hemodynamic parameters proposed previously. Third, our study populations had a broad age-range between 31 to 96 years old, not limited to aging population.

Two weakness were also noted in this study. Since our study was a cross-sectional design, the causality inference may be inappropriate. Further prospective studies need for elucidate this relationship. Second, the method to evaluate cognitive function in this study was MoCA, which was more sensitive than MMSE.^22^ Future external validation studies may be considered by using cognitive function assessment tools with different sensitivity and a wider distribution of cognitive function of the study population.

## Conclusion

Our study reveals that energetic hemodynamic parameters, especially carotid mean energy and carotid energy PI, provide a more robust framework for understanding the vascular-cognitive nexus compared to conventional measures.

### Perspectives

In summary, our research demonstrates a robust association between energetic hemodynamic parameters and cognitive function. There exists a strong association between the presence of an increased carotid energy PI, which is characterized by heightened carotid pulsatile energy and decreased carotid mean energy, and impaired cognitive performance, as assessed by the MoCA. Our study provides substantial evidence to support the notion that carotid energy PI and carotid mean energy are more dependable indicators of cognitive decline compared to conventional hemodynamic parameters that rely on flow or pressure measurements. This discovery affirms the enhanced explanatory potential of energetic hemodynamic parameters in the association between vascular health and cognitive function. Future emphasis may not be on arterial pressure or blood flow, but rather on energy, which may become the standard unit of measurement.

## Data Availability

The data can be made accessible upon reasonable request to the corresponding authors.

## Author contribution

Study Conception and designed the work: SY Chuang, HM Cheng, CH Chen, WH PAN, LK Chen, PN Wang, CP Chung, GF. Mitchell.

Bio-information extract: CH Lin and JJ Wang

Data analysis: SY Chuang and CJ Huang

Data Collection: SY Chuang, CH Chen, WH PAN, LK Chen, PN Wang, CP Chung.

Manuscript drafted: SY Chuang, HM Cheng, CH Chen, GF. Mitchell, JJ Wang

All authors have approved the submitted version.

## Novelty and Relevance

### What Is New?

Carotid energy PI is a significant indicator of cognitive health.
MOCA scores are more closely linked to energetic hemodynamics than conventional flow or pressure-based assessment.

### Relevance to Hypertension

Vascular aging plays a crucial role in the pathophysiology of hypertension.
This research suggests that energy-based hemodynamic parameters may better predict cognitive decline due to vascular aging in hypertension patients.

### Clinical/Pathophysiological Implications

Assessing the vascular-cognitive nexus may be more effective using carotid energetic hemodynamic parameters than pressure or flow-based frameworks.
Targeting hemodynamic changes may aid cognitive preservation strategies.
The study suggests a stronger link between vascular and cognitive health, which could inform future treatments for cognitive decline caused by vascular disease.

